# Towards a Universal Map of EEG: A semantic, low-dimensional manifold for EEG Classification, Clustering and Prognostication

**DOI:** 10.1101/2024.10.25.24316133

**Authors:** Laura Krumm, Dominik D. Kranz, Mustafa Halimeh, Alexander Nelde, Edilberto Amorim, Sahar Zafar, Jin Jing, Robert J. Thomas, M. Brandon Westover, Christian Meisel

**Author notes:** co-senior authors (M. Brandon Westover, Christian Meisel). Correspondence to: Christian Meisel, Charité – Universitätsmedizin Berlin, Comutational Neurology, Charitéplatz 1, 10117 Berlin, Germany.

## Abstract

Despite its complexity, whole-brain neural activity spontaneously organizes into a limited range of states, indicating that a low-dimensional representation or embedding might be sufficient to capture much of its macroscale dynamics. Clinical practice makes use of this notion by classifying EEG into discrete states, for example in the wake-sleep cycle or along the ictal-interictal continuum. Such classification into distinct classes has clinical value and simplifies EEG interpretation (by humans and, more recently, also by artificial intelligence [AI]) but misses subtle differences, mixtures and transitions between states. In other words, it does not adequately reflect the true underlying, low-dimensional continuum of states. The lack of a continuum EEG map limits more realistic characterizations of EEG states and challenges prognostication in patients with disorders of consciousness due to their diverse patterns, etiologies and underlying pathophysiologies. We here use data across the health-disease continuum (wake [W], sleep [REM, N1, N2, N3], ictal-interictal-continuum [lateralized and generalized periodic discharges (LPD, GPD), lateralized and generalized rhythmic delta activity (LRDA, GRDA)], seizures [SZ], burst suppression [BS]; 20,043 patients, 288,986 EEG segments) and a deep neural network that optimises a low-dimensional embedding space. This universal map of EEG (UM-EEG) is highly semantic, confined and captures meaningful distances as well as transitions between EEG states. Classification and clustering of unseen EEG are easily implemented using standard techniques on the map that match performance of current AI algorithms while extending it to the currently largest set of classes across the full health-disease continuum (mean AUROCs and 95% CI one-vs-all classification: W 0.94 [0.94, 0.95], N1 0.85 [0.85, 0.86], REM 0.92 [0.91, 0.92], N2 0.91 [0.90, 0.91], N3 0.98 [0.97, 0.98], GRDA [0.97 [0.96, 0.97], LRDA 0.97 [0.96, 0.97], SZ 0.87 [0.83, 0.91], GPD 0.99 [0.98, 0.90], LPD 0.97 [0.97, 0.97)], BS 0.94 [0.93, 0.94]). Long-term EEGs are represented as trajectories in this continuous embedding space which predict outcome (recovery or death) after cardiac arrest with an AUROC of 0.86 (*N* = 575 patients). For these patients, the map identifies interpretable factors governing prognosis: the distance to healthy states, the variability of state transitions, and time spent in burst suppression. These factors allow personalized monitoring and comparison of patients to themselves over time. In conclusion, UM-EEG presents a novel, comprehensive and physiologically meaningful representation to characterize and classify brain states along the health-disease continuum. It demonstrates immediate clinical utility by enhancing current diagnostic and prognostic capabilities.

## Introduction

The human brain spontaneously self-organizes into a limited continuum of states, such as those observed in the wake-sleep cycle, indicating that a low-dimensional representation or embedding might be sufficient to capture much of its macroscale dynamics.^1,2^ In mathematical terms, such low-dimensional representations are called manifolds, and are a key property characterizing the behaviour of many complex systems. The notion that manifolds capture important information from high-dimensional brain activity is supported by insights into the fundamental principles governing whole-brain neural activity patterns gained from low-dimensional representations, and their ability to dynamically track complex system behavior.^3–7^

The relevance and utility of low-dimensional brain state characterizations are further supported by the clinical use of electroencephalograms (EEGs) for classification into relatively few states and features.^8^ As a key diagnostic tool in neurology, EEG interpretation requires specialized expertise, which is often lacking,^9^ contributing to misinterpretation.^10,11^ To address the shortage of experts, increasing numbers of EEG referrals, and lack of expertise, artificial intelligence (AI) methods have been applied for automated classification of such states, including the detection of seizures, episodes of focal or global slowing, and identification of interictal epileptiform discharges.^12,13^ However, while classification according to distinct classes may simplify EEG reading and AI training, it does not adequately capture the underlying continuum of states, relationship and mixtures between states or their dynamical transitions. In other words: it does not adequately reflect the underlying manifold.

The challenge to appropriately capture individual trajectories within the health-disease continuum becomes particularly evident in long-term prognostication. Despite the availability of EEG, prognostication for patients with disorders of consciousness (DOCs) remains challenging due to their diverse etiologies and underlying pathophysiologies.^14^ Current outcome prediction methods primarily depend on expert multimodal evaluations of clinical variables and EEG, and typically involve visual assessments, which can be time-consuming and prone to subjective interpretation.^15^ Human visual interpretation may further be limited in appreciating complex long-term data trends, transitions, or mixtures of states, and may overlook subtle or not yet known features in EEG. Stimulation-based approaches for prognostication may require complex setups not readily available everywhere.^16,17^

Artificial intelligence has the potential to meet these challenges as it can, in principle, capture the full information contained in EEG, and represent it in low-dimensional manifolds. Previous AI models have focused on specific tasks, like distinguishing between normal and abnormal EEGs, local and global slowing or detecting epileptiform activity and seizures^12,13,18–20^. However, these binary approaches lack the ability to represent EEG as a health-disease continuum for truthful classification and delineation of time trajectories during individualized prognostication.

Here we leverage a large-scale EEG dataset across the health-disease continuum to derive a universal map of EEG (UM-EEG) within a compact Euclidean space where distances directly correspond to a measure of EEG similarity. The map exhibits a low-dimensional, semantic manifold that captures meaningful distances and transitions between brain states. This space allows for classification and clustering of out-of-sample data across a broad continuum of physiological and pathological states. For long-term data, it captures trajectories along the manifold, which provide individualized outcome prognostication after cardiac arrest as a first application. Finally, the embedding offers immediate and understandable insight into the factors governing prognosis after cardiac arrest, including the distance to healthy EEG dynamics, the variability of transitions between states or the time spent in burst suppression.

## Materials and Methods

### Datasets

To create a universal map of EEG we used data from multiple sources covering a broad range of physiological and pathological states: a sleep dataset from healthy subjects during polysomnography (PSG), a dataset covering the ictal-interictal-injury continuum (IIIC),^21^ data from awake routine EEG recordings,^22^ and burst suppression EEG data.^23,24^ We furthermore use continuous EEG data from patients diagnosed with a disorder of consciousness following cardiac arrest monitored over extended periods of time.^25–27^ Retrospective analysis of data for this project was conducted with waiver of informed consent under approved IRB protocols (BIDMC: 2022P000417; MGH: 2013P001024).

#### Polysomnography Data

The Human Sleep Project (HSP)^28^ dataset consists of clinical polysomnography (PSG) recordings of 25,941 patients, of which 3,609 patients were identified as medically healthy and included in our study.^29,30^ The PSG recordings included six channels of EEG (F3-M2, F4-M1, C3-M2, C4-M1, O1-M2, and O2-M1) based on the international 10-20 system. The dataset includes annotations of wake resting state and sleep stages: wakefulness (W), non-REM sleep stage 1-3 (N1-N3), rapid eye movement sleep (REM). Sleep stages were annotated in 30-second intervals by sleep technologists according to the American Academy of Sleep Medicine (AASM) manual for sleep scoring.

#### Ictal-Interictal-Injury Continuum Data

The Ictal-Interictal-Injury Continuum (IIIC) dataset includes EEG from 1,557 patients, recorded at Massachusetts General Hospital during clinical care.^21^ EEGs were referenced to a bipolar montage including the following channels: Fp1 - F7, Fp1 - F3, F7 - T3, F3 - C3, T3 - T5, C3 - P3, T5 - O1, P3 - O1, Fp2 - F8, Fp2 - F4, F8 - T4, F4 - C4, T4 - T6, C4 - P4, T6 - O2, P4 - O2 and were sampled at 200 Hz. In our study we included 58,117 10-second segments that had received at least 10 independent annotations from a group of 20 experts.^31^ Segments were categorized into the following groups: seizures (SZ), lateralized and generalized periodic discharges (LPD, GPD), lateralized and generalized rhythmic delta activity (LRDA, GRDA). We excluded the group ‘other’ (OTH, including all non-IIIC patterns) from further analysis. The labelled segments were pre-processed by applying a notch filter at 60 Hz and a band-pass filter between 0.5 and 40 Hz.

#### Awake Routine EEG Data

Awake resting-state EEG data was extracted from the Harvard Electroencephalography Database (HED) which encompasses data gathered from four hospitals: Massachusetts General Hospital (MGH), Brigham and Women’s Hospital (BWH), Beth Israel Deaconess Medical Center (BIDMC), and Boston Children’s Hospital (BCH).^22^ The database includes 164,707 EEG studies conducted on 65,167 patients. For our study, we selected 2,355 patients from the database with normal routine EEGs (19 EEG channels, 10-20 system). The purpose of including this dataset was to calibrate differences between the polysomnography and IIIC datasets when training our model. As the datasets originate from different sources with distinct electrode montages, recording devices and pathologies, we aimed to ensure that the model did not merely learn the differences in monitoring setups. The IIIC dataset is a subset of the HED, which also includes healthy resting state EEG data similar to that in the HSP dataset. To calibrate potential differences between the HSP and IIIC datasets, we extracted wake data from both the HSP and the HED and trained our model with wake data from both datasets under the same label (W).

#### Burst Suppression Data

The burst suppression dataset^23,24^ contains EEGs from 20 critically ill neurological patients from the ICU at Massachusetts General Hospital, recorded between August 2010 and March 2012. The recording durations are less than 90 minutes and consist of 19-channel EEGs according to the international 10-20 system. Two experienced clinical electroencephalographers manually annotated these EEGs as either burst or suppression. Definite burst or suppression epochs were defined as segments of the EEG where both reviewers agreed on the classification. Reviewers marked the beginning and end of all instances of “suppressions,” with all remaining EEG segments classified as non-suppressions or bursts.

#### Cardiac Arrest Data

We investigated long-term EEG data from 605 patients with disorders of consciousness due to cardiac arrest from seven academic hospitals in the U.S. and Europe within the International Cardiac Arrest REsearch consortium (I-CARE).^25–27^ These patients were admitted to the ICU after having a cardiac arrest with a return of heat function while remaining in a comatose state. EEG monitoring typically began within hours of cardiac arrest and continued for several hours to days, varying based on each patient’s condition. A continuous 19-channel EEG (10-20 system) was applied to all patients, although different channels are available for patients from different hospitals. For all patients the EEG recording start time is provided in hours after the cardiac arrest. Neurological outcomes were assessed 3 to 6 months after cardiac arrest. A good neurological outcome was defined as a Cerebral Performance Category (CPC) score of 1 or 2, indicating minimal to moderate neurologic disability. A poor outcome was defined as a CPC score of 3-5, indicating severe neurologic disability, persistent coma or vegetative state, or death. In this study, we only included patients with a good outcome of CPC 1 or 2 and a poor outcome of CPC 5 (in total 575 patients) as patients with outcomes 3 or 4 usually had large gaps between the end of the EEG recording and their final assessments.

### Data Preprocessing

We performed the following preprocessing steps to unify the different datasets. First, all data were resampled to 200 Hz, if necessary. A notch filter at 60 Hz was applied to remove line noise along with a 4^th^ order butterworth bandpass filter between 0.5 and 40 Hz. Next, all datasets were re-referenced to a common electrode configuration. The HSP dataset contained six EEG channels: F3-M2, F4-M1, C3-M2, C4-M1, O1-M2, and O2-M1, while the IIIC segments were already re-referenced as a bipolar montage (Fp1 - F7, Fp1 - F3, F7 - T3, F3 - C3, T3 - T5, C3 - P3, T5 - O1, P3 - O1, Fp2 - F8, Fp2 - F4, F8 - T4, F4 - C4, T4 - T6, C4 - P4, T6 - O2, P4 - O2). To unify across datasets, all EEGs were re-referenced to the common configuration: F3 - C3, C3 - O1, F4 - C4, C4 - O2 (Fig. 1A). Since the IIIC dataset had been segmented into 10-second intervals already, non-overlapping 10-second segments were extracted from all other datasets as well. For the sleep data, annotations were provided for 30-second increments of which we extracted the middle 10 seconds.

**Figure 1.**
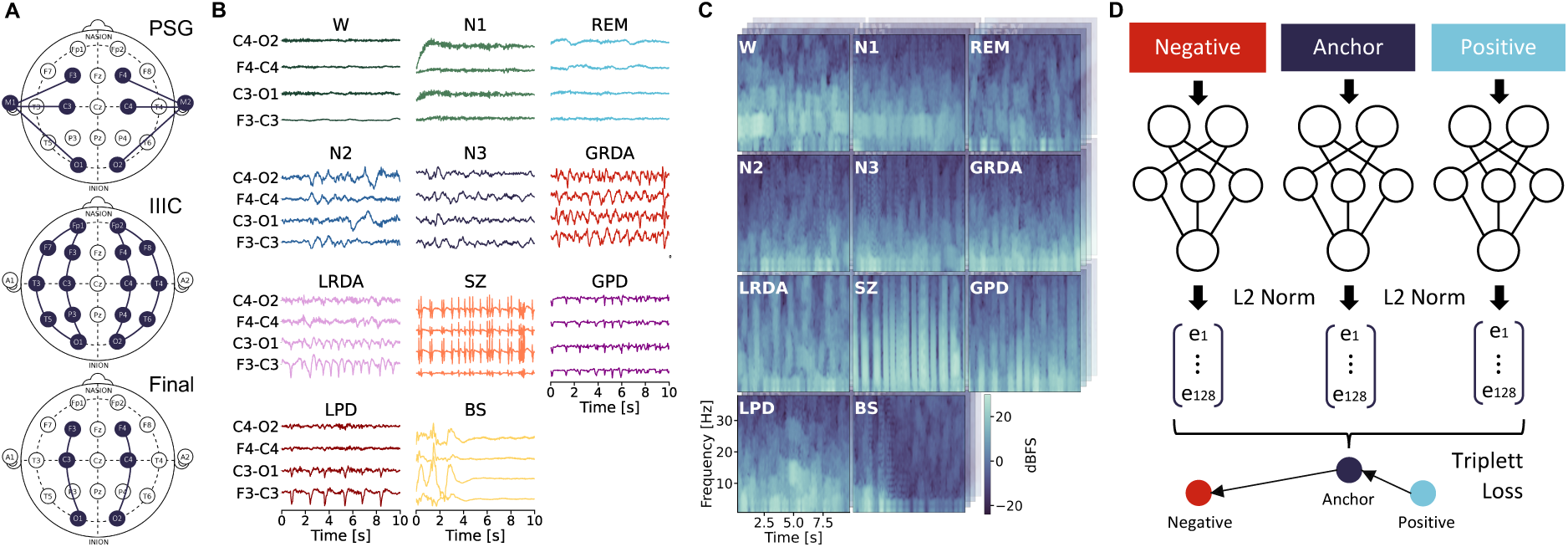
Data, pre-processing and deep learning model. (**A)** EEG re-referencing to include polysomnography (PSG) and Ictal-Interictal Injury Continuum (IIIC) data. (**B**) Example segments for different activities (W (wake), REM (rapid eye movement sleep), non-REM sleep stages N1-N3, SZ (seizures), GRDA (generalised rhythmic delta activity), LRDA (lateralised rhythmic delta activity), GPD (generalized periodic discharges), LPD (lateralised periodic discharges) and BS (burst suppression). (**C**) Corresponding spectrograms, channel C4-O2 in the front. (**D**) Schematic representation of the model and training in triplets.

The IIIC dataset comprised an average of 1,557 patients, with approximately 58,117 segments per class (SZ, GPD, LPD, GRDA, LRDA). For the sleep data, we utilized EEGs from 3,609 patients. To maintain a comparable magnitude of segments to the IIIC classes, we employed a systematic sampling approach. We first determined the number of available segments for each patient across all sleep stages and the wake state. Subsequently, we extracted either the maximum number of available segments or up to 10 segments per class, whichever was lower. This method ensured a balanced representation across sleep stages while preventing oversampling from any single patient or state. For the routine EEG data, 10 segments (each 10-seconds long) were randomly chosen and extracted per patient. For the burst suppression data, 10-second segments in which both experts annotated at least 50% of the time as suppression periods were selected, with a maximum of 2,000 segments per patient or as many as possible, if less than 2,000 segments were available. Due to the limited number of patients (20 patient EEGs), more segments were extracted for each patient here, compared to the other datasets. For the cardiac arrest data, segments were extracted in consecutive, non-overlapping 10-second steps across the full recording duration.

Finally, each 10-second segment from every EEG channel’s signal was converted to a spectrogram (multitaper method^32^ with time bandwidth = 2, number of tapers = 3, window parameters = [0.5, 0.1] min, nfft = 512, detrend option = constant, multiprocess = True, weighting = ‘unity’) resulting in four spectrograms per each 10-second data segment (Fig. 1C).

### Model Development

We used EEG spectrograms to develop a comprehensive latent space representation across physiological and pathological brain states. Segments (i.e. spectrograms) were split into training and test sets at the patient level using an 80/20 ratio, ensuring that all segments from any given patient appeared only in either the training or test set. Both sets thus contained a similar proportion of EEG segments for the different classes. To train a latent space EEG map, we adopted an approach based on triplet loss learning that has been successfully used in domains such as face recognition and clustering.^33^ The model maps data into a compact, 128-dimensional Euclidean space, where the embeddings are projected onto a unit hypersphere, ensuring they lie within a confined space. A triplet loss function optimizes the network weights during training, by minimizing the distance between embeddings of the same class and maximizing the distance between embeddings of different classes (Fig. 1D). More specifically, the network processes three 10-second segments (each consisting of 4 spectrograms for the 4 EEG channels, respectively) simultaneously: an anchor (a segment from a certain class, e.g., W), a positive (another segment from the same class, e.g., W), and a negative (a segment from a different class, e.g., N1). The model architecture, a Siamese Inception network, shares weights across the three branches and updates them simultaneously. This design ensures that the learned embeddings accurately reflect the similarity between inputs, creating a space where distances correspond to similarity.^33^ To train the model with triplet loss, we instantiated a data generator that selects anchor, positive, and negative segment for each training batch. The generator randomly chooses one of the 11 training classes (W, N1, N2, N3, REM, SZ, GPD, LPD, GRDA, LRDA, BS) as the anchor class. It then randomly selects an anchor segment and a positive segment from the chosen class. A negative class is selected at random from the remaining classes, and a segment from this class is fetched. This approach ensures that all class combinations are explored during training, promoting robust learning of class-specific features. The batch-based selection process enhances efficiency and ensures comprehensive coverage of the dataset.

### Performance Evaluation

To evaluate the model’s performance, we monitored the loss function throughout the training process and made sure that the loss was not decreasing any longer before stopping. Additionally, we generated embeddings for a subset of the test data, consisting of 1,000 spectrograms per class, at specific epochs (epochs 0-5 in increments of 1, and epochs 10-50 in increments of 10) and trained a support vector machine (SVM) using these test embeddings of the 11 classes (W, N1, N2, N3, REM, SZ, GPD, LPD, GRDA, LRDA, BS)to monitor the F1 score for clustering of unseen data over time. After completing training, we saved the final model and generated embeddings for the complete test dataset. To evaluate the final model’s ability to cluster unseen data, we used 5-fold cross-validation (at the patient level) using a support vector machine. Additionally, we computed the confusion matrix for all five folds and reported the mean and standard deviation for each entry. We also performed a binarized classification analysis by computing the area under the receiver-operating-characteristic curve (AUROC) and under the precision-recall curve (AUPR) for each class against all others (one-vs-all). For each fold, we applied bootstrapping (50 iterations per fold) on the test-fold data to calculate the mean ROC, PR curves, respective AUC values, and their 95% confidence intervals.

To assess the semantic nature of the 128-dimensional embedding space, we utilized multidimensional scaling (MDS), a statistical technique that visualizes the level of similarity or dissimilarity between a set of objects in a lower-dimensional space. We first calculated the median embeddings for each class in the 128-dimensional embedding space and then computed a distance matrix using cosine distances between all class medians. We applied MDS to this distance matrix to reduce the dimensionality to 2D, allowing us to visualize the relative positions of the class medians in a two-dimensional space (Fig. 2). To better comprehend the spatial distribution of our burst suppression data in 128-dimensional space, we calculated the Burst Suppression Ratio (BSR) for each burst suppression segment. The BSR, also known as the Burst Suppression Index (BSI), was computed using the following parameters: maximum amplitude of suppression of 10 microvolts, minimum duration of suppression of 0.5 seconds, and minimum duration of bursts of 0.2 seconds. We then investigated the spatial arrangement of burst suppression patterns within the map across three categories: high proportion of suppression periods (BSR > 0.7), intermediate proportion of suppression and burst periods (0.35 ≤ BSR ≤ 0.65), and high proportion of burst periods (BSR < 0.3).

**Figure 2.**
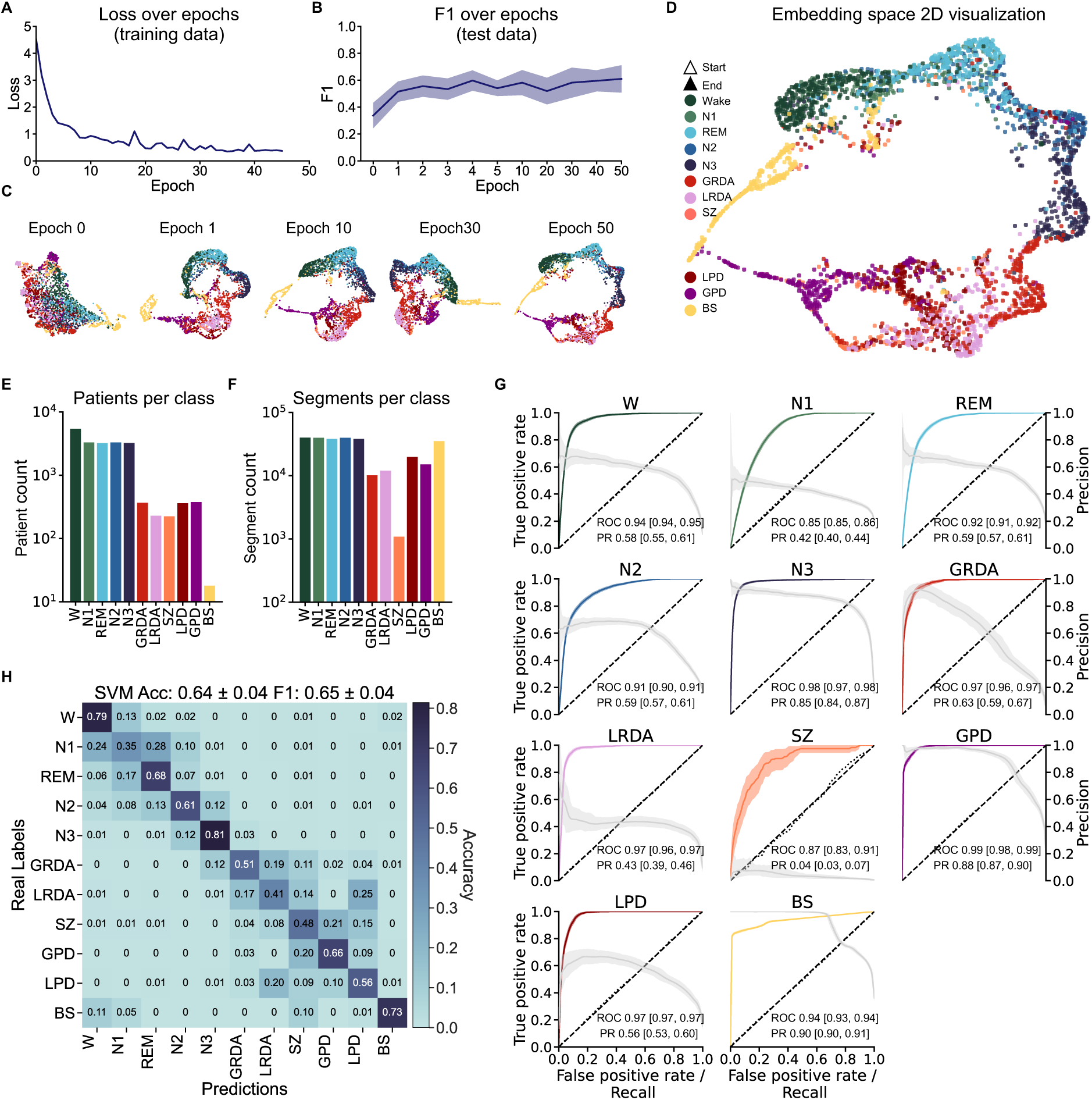
Training and state classification using the embedding space. **(A)** Loss over epochs during training. (**B)** For selected epochs, the same 1000 test segments per class were passed through the model and classification performance was monitored with F1 scores (support vector machine (SVM), 5-fold cross-validation). **(C)** Embedding space of 300 test data points in 2D representation (dimensionality reduction via UMAP) for the selected epochs. (**D)** Final embedding space representation in 2D via UMAP. (**E)** Number of patients per class for all data (80% training, 20% testing data). **(F)** Number of 10-sec segments per class for all data. **(G)** ROC and PR curves for a SVM trained on embeddings from the test set via 5-fold cross-validation. For each fold bootstrapping (50 times) was performed on the test data to compute the mean ROC / PR curves and their respective AUCs, as well as their 95% confidence intervals. (**H)** Confusion matrix for the mean classification accuracy between the test embeddings of the different classes using a SVM.

### Prognostication of Patient Outcomes from Trajectories in Embedding Space

We projected continuous data from post-cardiac arrest comatose patients into the universal map of EEG to obtain trajectories in latent space. Specifically, for each patient, we processed consecutive 10-second EEG segments through the map, generating a time series of embedding vectors. To assess the predictive power of these embedding trajectories for patient outcomes, we first projected the EEG into either one of the 11 classes (W, N1, N2, N3, REM, SZ, GPD, LPD, GRDA, LRDA, BS) by calculating cosine distances from each time point to the median coordinate of each of the 11 classes. We then assigned each segment to the nearest class to analyse how patients with different outcomes traversed the embedding space. Next, we quantified for each patient the relative time spent in each of the 11 classes. We then used five-fold cross-validation using a Support Vector Machine (SVM) to classify patients according to their outcomes (CPC 1 and 2, i.e. recovery, vs. CPC 5, i.e. death). We report performance in terms of AUROC and the corresponding 95% confidence intervals.

Recognizing that our map was trained on specific EEG patterns and that the cardiac arrest dataset might present a spectrum of patterns not directly aligning with the predefined classes used in training, we also conducted a more granular analysis of patient trajectories. Specifically, we generated an evenly spaced grid on the unit hypersphere by randomly sampling up to 100,000 data points in 128 dimensions, projecting them onto the hypersphere, and applying repulsive forces to distribute them evenly across the surface. Each point was then assigned a reference number. We then computed the cosine distance from each patient data point to the nearest grid point rather than the nearest class median. This approach allowed for a finer resolution of the trajectories within the continuum embedding space, providing a detailed mapping of patient states without strict reliance on the predefined classes.

### Dynamics of Trajectories and Linear Discriminant Analysis

To assess the dynamical information contained in the trajectories in our map, we calculated 45 parameters from classical time series analysis (Supplementary Table 1). These parameters included temporal and symbolic dynamics as well as complexity measures derived from the time series of classes (W, N1, N2, N3, REM, SZ, GPD, LPD, GRDA, LRDA, BS), as well as spatial measures derived directly from the 128-dimensional embeddings. We derived these parameters from the time series of classes in the cardiac arrest dataset. We investigated which combinations of two or three parameters best separated the two outcome groups (CPC 1 and 2 vs. CPC 5) by performing a linear discriminant analysis (LDA) for each combination of parameters. This approach demonstrates the utility of the embedding to derive explainable and intuitively understandable dynamics parameters that adequately predict neurologic outcome.

## Results

### Building a Universal Map of EEG (UM-EEG)

We included EEGs from a wide range of physiological and pathophysiological states, including the healthy / normal brain activity continuum (W, N1, N2, N3, REM), the ictal-interictal-injury continuum (GPD, LPD, GRDA, LRDA, SZ) and burst suppression (BS), to create a comprehensive latent space map of EEG (Fig. 1). Systematic initial evaluations on the Ictal-Interictal-Injury Continuum (IIIC) dataset indicated that performance increased strongly when more than one channel was used and then reached some stable plateau around four channels with only a minor performance increase when all 16 channels were used (Supplementary Fig. 1). We thus re-referenced all data to four bipolar channel pairs to include data from all datasets (Fig. 1A). Model performance monitored with loss function and F1 score indicated stable performance improvement during training without signs of overfitting (Fig. 2A, B). For visualization purposes, the 128-dimensional embedding test data was projected onto a 2-dimensional space using Uniform Manifold Approximation and Projection (UMAP).^34^ As training progressed, the classes exhibited increased clustering, ultimately resulting in a complex embedding space with some classes disjunct from and others merging into each other (Fig. 2D). Notably, already in the 2D projection, semantic clustering emerged, including distinct and meaningful spatial arrangements capturing, for example, natural state transitions during sleep (W → N1 → REM → N2 → N3).

### Classification of unseen EEG in UM-EEG

We first assessed classification of EEGs in the 128-dimensional embedding space using the receiver-operating-characteristic (ROC) and precision-recall (PR) curves for all classes in a binarized classification approach (one-vs-all; Fig. 2G). The mean area under the ROC curve and corresponding 95% confidence intervals for each class indicated robust class separation: W (0.94 [0.94, 0.95]), REM (0.92 [0.91, 0.92]), N1 (0.85 [0.85, 0.86]), N2 (0.91 [0.90, 0.91]), N3 (0.98 [0.97, 0.98]), GRDA ( [0.97 [0.96, 0.97]), LRDA (0.97 [0.96, 0.97]), SZ (0.87 [0.83, 0.91]), GPD (0.99 [0.98, 0.90]), LPD (0.97 [0.97, 0.97)]) and BS (0.94 [0.93, 0.94]). For the IIIC (SZ, LPD, GPD, LRDA, GRDA), apart from seizures, these results match or exceed discrimination to recently published results,^21^ while including more classes. The relatively lower performance for seizures (SZ) may be attributed to the limited number of segments available for this class and the high variability of patterns within this category. Similarly, AUPR curves showed effective binary classification based on the latent space map (Fig. 2G). Across all 11 classes, the model demonstrated effective learning in clustering achieving a final F1 score of 0.65 ± 0.04 (compared to 0.08 ± 0.01 for a random predictor) and an accuracy of 0.64 ± 0.04 (compared to 0.09 ± 0.01 for a random predictor) for multiclass classification. Fig. 2H shows the confusion matrix across all individual classes. Collectively, these results demonstrate effective classification of out-of-sample data based on distinct clustering in latent space. This suggests a highly semantic embedding space of the various EEG states and patterns.

### Semantic representation across the health-disease continuum in UM-EEG

In a semantic latent space, similar data points are positioned closer together, reflecting their semantic (meaningful) relationships. To determine semantic properties of the EEG map beyond the 2D UMAP visualization, we next analysed the 128-dimensional embedding space. Specifically, we computed the median vectors for all classes in this space and calculated the cosine distances between class medians. Additionally, multidimensional scaling (MDS) was employed to create a 2D representation that preserves the inter-class distances from the 128-dimensional space as closely as possible (Fig. 3A). For a precise comparison, we also presented all pairwise distances in a matrix format (Fig. 3B). Results from this analysis revealed a meaningful spatial arrangement in the 128-dimensional space, consistent with MDS and visual assessment of the 2D UMAP projection.

**Figure 3.**
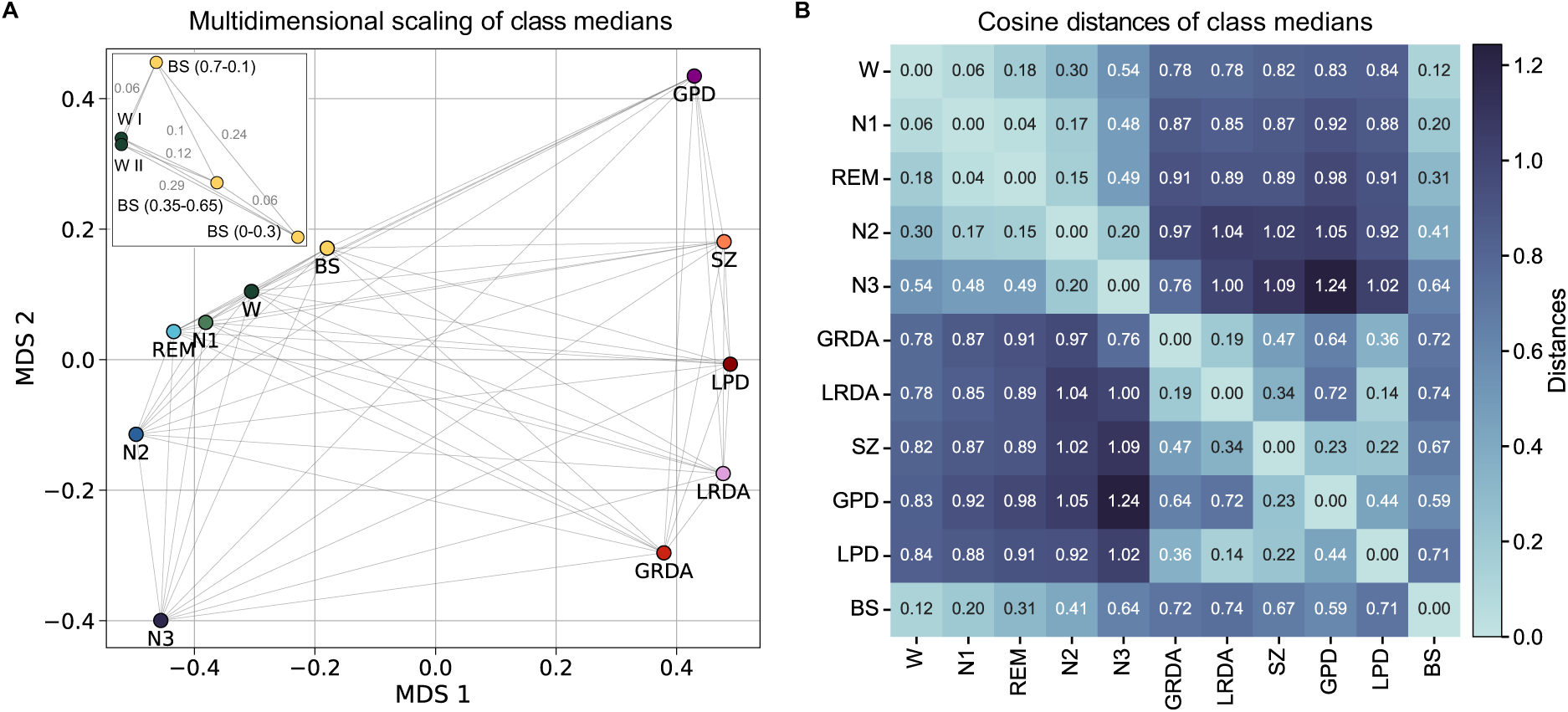
Semantic representation of EEG states across the health-disease continuum in embedding space. **(A)** Class medians and their respective cosine distances are projected into 2D using multidimensional scaling which preserves the spatial arrangement in 128D as closely as possible. The inlay provides a more nuanced distinction within the burst suppression and wake classes. Our wake resting state data originates from two distinct datasets (WI from the HSP and WII from the HED dataset). For the burst suppression data, we applied an additional grouping based on the Burst Suppression Ratio (BSR). This grouping resulted in three categories: high proportion of suppression periods (BSR > 0.7), intermediate proportion of suppression and burst periods (0.35 ≤ BSR ≤ 0.65), and high proportion of burst periods (BSR < 0.3). (**B)** Matrix representation of the cosine distances of all class medians in 128D.

First, healthy and pathological states appeared to be clearly separated. The pathological domain (GPD, SZ, LPD, LRDA, GRDA) was far from physiological states, with generalized rhythmic and periodic patterns (GPD, GRDA) flanking the respective lateralized analogues (LPD, LRDA). Second, arrangement within the physiological domain closely reflected the natural state transitions of sleep stages within sleep cycles (W → [N1, REM] → N2 → N3). Third, the model captured nuanced differences within the BS category and its relationship to normal wake patterns. Specifically, in a subanalysis, we computed the mean Burst Suppression Ratio (BSR) across all channels from BS segments to quantify the proportion of burst and suppression patterns more precisely. We observed that EEG segments with BSR < 0.3 (high proportion of suppression) were more distant from wake (cosine distance = 0.29). Patterns with BSR between 0.35 and 0.65 showed an intermediate distance to wake (cosine distance = 0.12) whereas patterns with BSR between 0.7 and 1 (burst-dominant) were closest to wake (cosine distance = 0.06; Fig. 3A inset). These results indicate that there is a gradient in the embedding space, where BS patterns with a higher proportion of bursts are arranged closer to the wake state, and, conversely, BS patterns with higher proportion of suppression are arranged further away from wake. Fourth, the model correctly assigned wake (W) EEG from different datasets to the same place in the map. As our W data class was comprised from two distinct datasets (HSP wake data and HED resting state), we projected both W test data sources (W I and W II) separately into the map in a subanalysis (Fig. 3A inset). We found the class medians for these two to be in very close proximity (distance = 0), indicating no significant differences between the datasets, and that, consequently, distances between states could not simply be attributed to originating from different datasets.

### Patient trajectories in UM-EEG predict outcome after cardiac arrest

We hypothesized that projections of long-term EEGs into the semantic embedding would allow for effective characterization of these trajectories relevant for outcome prediction and identification of important predictive data signatures. We generated patient trajectories from 575 patients after cardiac arrest by feeding consecutive 10-second segments into the model, mapping each patient’s EEG evolution over time in embedding space (Fig. 4A, B).

**Figure 4.**
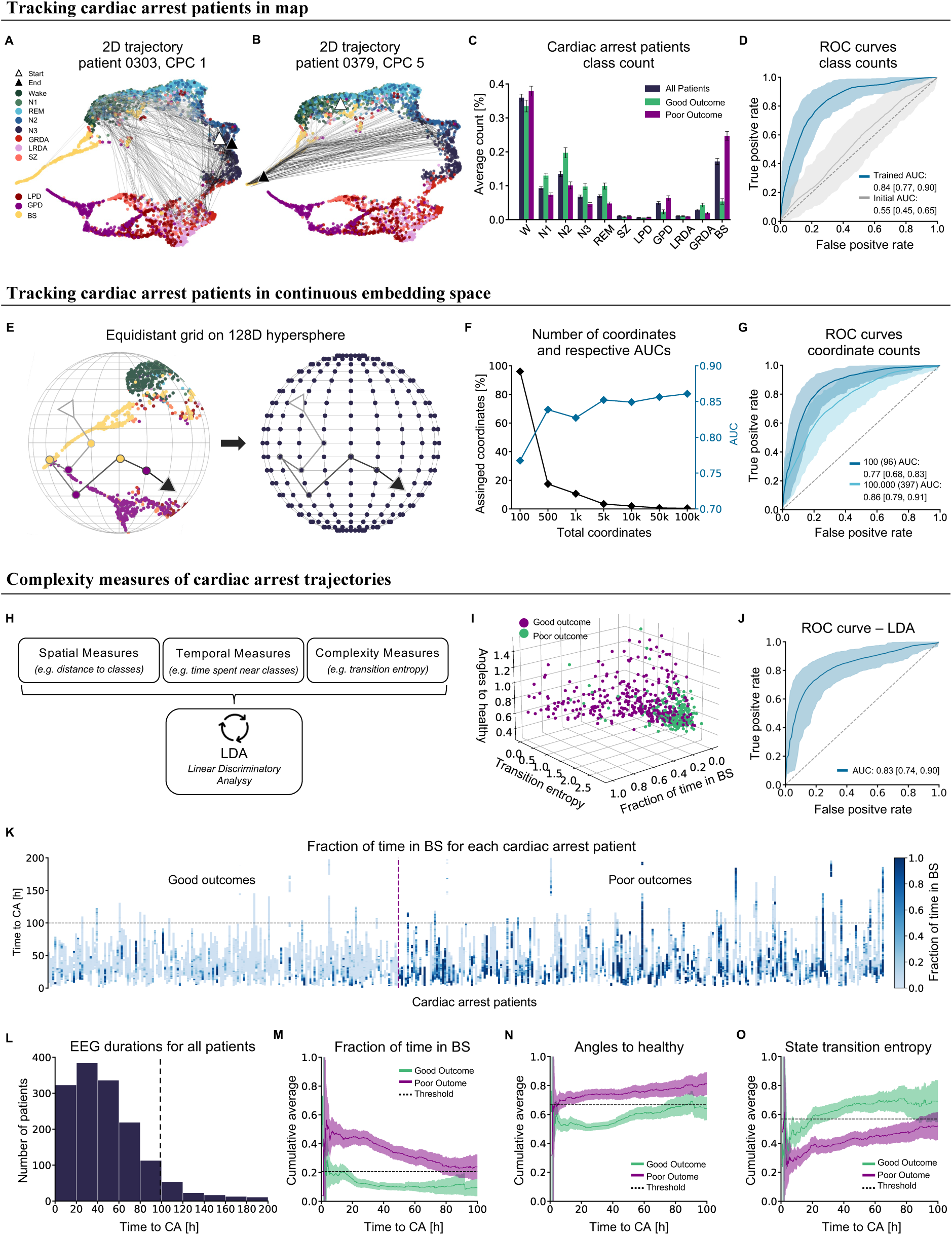
Patient trajectories in UM-EEG predict outcome after cardiac arrest. (**A**) 2D UMAP representation (of 128D hypersphere) for a cardiac arrest patient trajectory in with a good outcome (CPC 1) projected into the map over time. The white triangle indicates the starting point and the back triangle the end of the trajectory. (**B**) Example trajectory of a cardiac arrest patient with a poor outcome (CPC 5). (**C**) Average time spent in each class according to outcome. Each trajectory segment was assigned to the class with the smallest cosine distance. The class counts for all patients were normalized by their total number of segments and averaged. The error bars indicate the standard error of the mean. (**D**) ROC curves for classification of good (CPC 1,2) vs poor (CPC 5) outcome based on class counts for each patient in trained (blue) vs untrained embedding space (grey). (**E)** Generation of an equidistant grid (coordinates) on the 128-dimensional hypersphere. Instead of tracking cardiac arrest patients in terms of closest classes, they were assigned to the closest coordinates for finer resolution. (**F)** Performance as a function of total size of coordinate system and the percentage of closest coordinates. **(G**) ROC curves for cardiac arrest outcome prediction on grids of 100 and 100000 coordinates. (**H)** Schematic overview of metrics used for time series analysis of trajectories. (**I)** Visualization of separation of outcome achieved with three best linear discriminant analysis (LDA) parameters (time spent in burst suppression, distance to healthy and state transition entropy). (**J)** Corresponding ROC curve. (**K)** Individual patient trajectories according to time spent in burst suppression. (**L)** Histogram of durations of EEG data. We chose a cut-off after 100 hours due to insufficient data. (**M, N, O)** LDA parameters provide intuitive markers predictive of prognosis after cardiac arrest. Dashed horizontal lines indicate the center between the mean of patients with good outcomes and the mean of patients with poor outcomes.

We first assessed the prognostic information content contained in map trajectories by assigning each 10-second segment to the 11 training classes according to the smallest cosine distance to class medians (Fig. 4A, B). The relative time spent in different classes differed between patients with good (CP 1 and 2) and poor outcomes (CPC 5, Fig. 4C). Patients with good outcomes spent more time in states close to healthy conditions (W, N1, N2, N3, REM) compared to those with poor outcomes (*P* = 1.99e-16, Mann-Whitney U test). Conversely, patients with poor outcomes spent more time in pathological states (SZ, LPD, GPD, LRDA, GRDA; *P* = 2.29e-02) and particularly in burst suppression (*P* = 1.47e-27). Using the information of relative time spent in these classes provided good outcome prediction after cardiac arrest with an AUROC of 0.84 [95% CI: 0.77, 0.90] (Fig. 4D). In comparison, projections into an untrained map yielded a significantly worse AUROC of only 0.55 [95% CI: 0.45, 0.65] (*P* = 2.19e-224), demonstrating that the embedding space, trained on both healthy and pathological classes, enhanced the meaningfulness and prognostic value of cardiac arrest EEGs.

Assigning segments to the nearest training class provided already good prediction performance result but effectively transformed the problem into a classification task limited by the number of classes the embedding was trained on. Given the highly semantic nature of the embedding space and the heterogeneity of cardiac arrest EEG patterns, we hypothesized that outcome prediction would improve further when trajectories were not limited to predefined classes but could make use of the full continuum spanned by our map. We therefore applied a more nuanced approach that leveraged the continuous nature of the embedding space by projecting an evenly spaced 128-dimensional grid onto this sphere to serve as a coordinate system (Fig. 4E). Then, instead of assigning each segment merely to the closest class, we assigned it to the closest coordinate. As we increased the number of coordinates, making the grid more fine-grained, the AUROC increased (Fig. 4F) leading to an AUROC of 0.86 [95% CI: 0.79, 0.91] for the most fine-grained coordinate system with 100,000 coordinates (from 0.77 [95% CI: 0.68, 0.83] with 100 coordinates, *P* = 4.38e-72). This continuum-based approach outperformed prognostication based on traditional classes (*P* = 1.40e-07, Fig. 4G). While increasing the number of coordinates improved the AUROC, the number of coordinates actually used increased only slightly (96 coordinates used for a grid size 100, 397 for grid size 100,000; Fig. 4F). This indicates that the AUROC improved due to a finer grid representation of the cardiac arrest data clustered in distinct areas across the hypersphere.

Finally, we determined what dynamical characteristics of the trajectories in embedding space, aside from the relative time spent in certain states, were predictive of outcome. For this purpose, we derived well established spatial, temporal and complexity parameters from the time series of classes, as well as the trajectories directly (Supplementary Table 1) and applied Linear Discriminant Analysis (LDA) to identify parameters that separated “poor” and “good” outcomes (Fig. 4H). The best discrimination was achieved with three parameters, indicating that a low distance (angle) to the healthy continuum together with a low amount of time in burst suppression and a high state transition entropy were predictive of a good outcome (Fig. 4I). While using only these three parameters provided a comparable AUROC (0.83 [95% CI: 0.74, 0.90], Fig. 4J), the simplicity of these parameters exemplifies the predictive, meaningful and understandable information gained from the map that can be used for monitoring patients. Specifically, these metrics match the intuitive assumption that a healthy brain displays complex patterns in state transitions, is closer to the healthy states (wake and sleep) and spends little to no time in burst suppression. Meaningful separation in these metrics was already evident after the first 12 hours after cardiac arrest (Fig. 4M, N, O) indicating that these metrics can be used to benchmark a patient to themselves over time and, potentially, also in other, heterogenous diseases and patient cohorts beyond cardiac arrest.

## Discussion

We here present a latent space embedding for EEG that covers the health-disease continuum and preserves the physiologic ordering within brain states. Our findings indicate that various brain states, physiological and pathological ones, can be represented and understood as points or trajectories within this low-dimensional latent space, offering new applications in diagnosis, prognosis, and data augmentation for automated classification.

To generate our universal map of EEG, we projected the training data onto a confined space (a 128D hypersphere) while allowing the data to arrange itself meaningfully. Perhaps surprisingly, we found that data from the Inter-Ictal-Injury continuum can be accurately represented in our embedding space despite a lower channel count (4 channels instead of 16). This enabled the creation of a unified space that incorporates diverse datasets, including polysomnography data where only a low channel count is typically available. The resulting embedding space not only organizes the training classes in a semantically significant manner but also enables a continuum between different classes. This continuum reflects, for example, the natural progression of sleep stages (W → [N1, REM] → N2 → N3), suggesting that the map captures a lower-dimensional manifold of sleep stage transitions within this higher-dimensional space. Moreover, pathological states are arranged much further away from wake than other healthy states (sleep stages). The continuous and semantic nature of this embedding space offers an additional advantage: it allows for the meaningful projection of unseen data that may not precisely match the training classes. New data self-arranges within the space in semantically appropriate positions, demonstrating flexibility with novel inputs. Our approach thus not only demonstrates the potential for complex data relationships to be represented in a more interpretable form but also provides a framework for categorizing unseen data.

Although the model is not a classifier per design, classification naturally emerges from our embedding space as a spatial clustering problem that can be solved using standard clustering algorithms such as SVMs. Framing classification as a clustering problem enables us to easily classify both physiological and pathological EEG patterns, which exceed current AI approaches which are mostly limited to only a few pathological classes.^12,13^ Therefore, direct comparison of our study to existing research is challenging. For classes derived from the IIIC dataset, our method matches or exceeds the state of the art, for 4 out of 5 AUROC and 2 out of 5 AUPR scores (AUROC comparison of our results with state of the art: SZ 0.87 vs 0.92, LPD 0.97 vs 0.96, GPD 0.99 vs 0.93, LRDA 0.97 vs 0.94, GRDA 0.97 vs 0.80 and AUPR comparison: SZ 0.04 vs 0.78, LPD 0.56 vs 0.91, GPD 0.88 vs 0.75, LRDA 0.43 vs 0.70, GRDA 0.63 vs 0.48).^21^ Importantly, while a classification approach yields valuable insights, it may not adequately capture the full complexity of EEG patterns. In this study we showed that states often exist on a continuum, so reducing analysis to discrete classification results in some loss of information.

As a first use case, we projected data from cardiac arrest patients into the embedding space to predict outcome. Our approach (AUROC 0.86 [95%CI 0.79, 0.91]) surpassed existing stimulation-based tests in predicting outcomes for cardiac arrest patients using only resting-state EEG data (AUROC 0.70 ± 0.04).^16^ Our prognostication results improved further when using an approach that captured more continuous information compared to a classification approach (AUROC of our grid compared to map classification). While our predictive performance was lower than a specifically trained classifier for the same dataset (AUROC 0.91 [95%CI 0.88-0.93]),^35^ the strength of our approach lies in its flexibility and generalizability. Unlike cohort-specific methods, that are common in machine learning, we suggest that our approach can handle patients with additional diseases or atypical cardiac arrest progressions. Moreover, our method is applicable across different etiologies without the need for retraining the model. As we predict outcomes based on a reduced number of interpretable measures such as distance to healthy state, time spent in burst suppression, and state transition entropy, our approach still provides good prognostic value (AUROC 0.83) while offering generalizability and interpretability. We provide meaningful scores for clinicians that could be monitored over time and aid in decision-making. Providing interpretable scores also offers the opportunity for individualized assessments. Specifically, we here demonstrate capability to generate personalized trajectories for each patient within the embedding space that can be transformed into meaningful information by applying time series metrics that would not be applicable to raw EEG. This approach thus allows patients to be benchmarked against their own baselines over time rather than against other patients in the same cohort, as is commonly practiced in machine learning.

Further use cases of UM-EEG might include clustering patients to relate them to similar individuals with existing medical reports, thereby aiding clinicians in their diagnostic processes and EEG reading. Additionally, it could enable the generation of new reports based on those of spatially close patients within our embedding space using large language models (LLMs). Beyond this, our universal map of EEG can have several meaningful clinical applications. Even in countries with advanced healthcare systems, many EEGs are interpreted by physicians who lack specialized fellowship training in this area, and which also contributes to misinterpretation of EEGs.^10,11^ Furthermore, the growing number of EEG referrals has created a significant workload, even for specialized centers, a problem which is particularly exacerbated during interpretation of long-term EEGs, including prognostication. Our approach could be implemented at the bedside as an end-to-end pipeline, offering automated patient monitoring and serving as a decision support system for clinicians. We hope to contribute to enhancing patient care by providing real-time, data-driven insights to healthcare professionals, potentially improving treatment efficacy. To work towards this goal we will continue training, expanding and validating UM-EEG.

While our approach demonstrates promising results, it is important to acknowledge its limitations. Our current use of only four EEG channels may result in the loss of important information, particularly for localized patterns such as activities from the Inter-Ictal-Injury-Continuum (LRDA and LDA). However, our subanalysis revealed that we still achieve good classification performance with this limited channel count compared to a higher channel count of 16 electrodes, suggesting the robustness of our method. As the model is trained in a supervised manner, another consideration is the potential for inaccuracies in our labelled segments (given they were labelled by humans). However, our embedding space is a continuum, which allows for more flexible placement of ambiguous or potentially mislabelled segments compared to a traditional classifier. Another limitation could be that our EEG segments are converted into spectrograms and future work will determine the utility of other data representations.

In conclusion, we here present, for the first time, a universal map of EEG as a comprehensive, highly semantic and confined embedding space that captures physiologically meaningful distances and transitions between EEG states. As a unified embedding space, UM-EEG affords characterization and classification of brain states in a nuanced way across the health-disease continuum and enhances current diagnostic and prognostic capabilities. The map provides opportunity for further growth and expansion and opens new avenues for personalized patient monitoring and data-driven clinical decision support.

## Data Availability

All data produced in the present study are available upon reasonable request to the authors.

## Supplementary Material

**Supplementary Figure 1.**
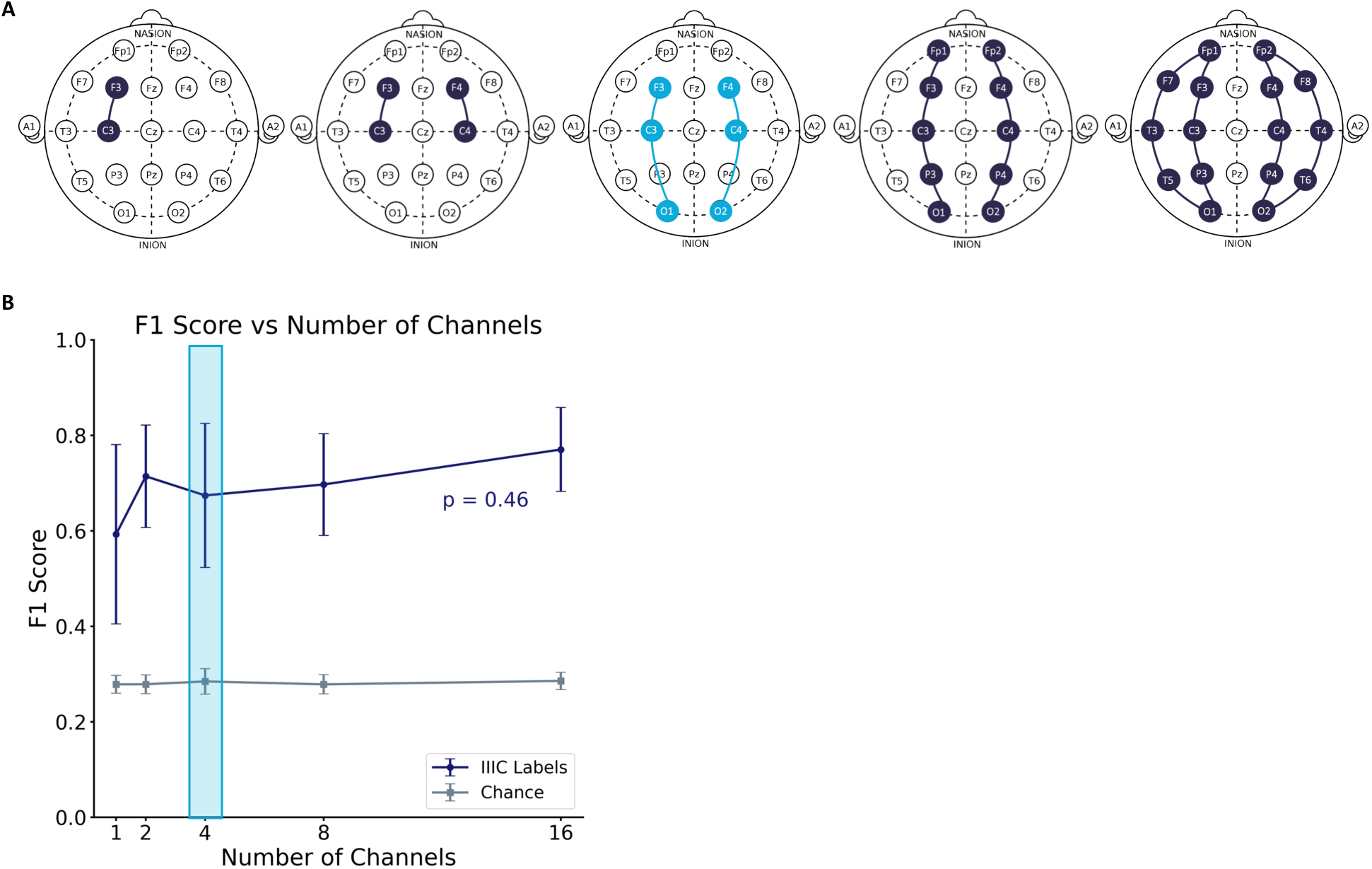
Map of IIIC data with different electrode configurations. **(A)** Illustration of EEG channels of IIIC data selected for training. The electrode configuration with 4 channels (light blue) represents the setup used for creating UM-EEG. **(B)** F1 score after training the model for the different electrode configurations on IIIC data. We performed an ANOVA test on the F1 values of the different electrode configurations (p-value: 0.46). The grey plot indicates F1 scores for an SVM trained on our test embeddings with shuffled labels (chance).

**Supplementary Table 1.** Parameters for time series analysis. Symbols denote: a – wake, b - N1, c - N2, d - N3, e – REM, f – SZ, g – LPD, h – GPD, I – LRDA, j – GRDA, k – BS. Words consist of three symbols. Consequently, a word could, for example, be: "aab" or "kak". From these words, we obtain a new time series, e.g. aab kak dff eee eff. All word parameters are then calculated on this time series.

|  |  |
| --- | --- |
| fwshannon | This parameter takes the histogram of words for a patient's whole recordings, and calculates the shannon entropy (= content of information) for them |
| forbword | Number of words that occur less than 0.01% of the time |
| healthyword | fraction of words that only contain healthy symbols (a, b, c, d, e) |
| deltaword | fraction of words that only consist of the symbol I and j |
| dischargeword | fraction of words that only consist of the symbols g and h |
| bsword | fraction of words that only consist of the symbols g and h |
| sleepword | fraction of words that only consist of the symbols b, c, d, e |
| deepsleepword | fraction of words that only consist of the symbol d |
| remsleepword | fraction of words that only consist of the symbol e |
| entropy_of_healthy_states | Shannon entropy of the symbol distribution for healthy symbols (a, b, c, d, e) |
| entropy_of_unhealthy_states | Shannon entropy of the symbol distribution for pathological symbols (f, g, h, I, j, k) |
| pathological_to_total | fraction of symbols that are pathological |
| healthy_to_total | fraction of symbols that are healthy (redundant, 1 - pathological) |
| longest_run_of_a | the longest repetition of the symbol a |
| longest_run_of_b | the longest repetition of the symbol b |
| longest_run_of_c | the longest repetition of the symbol c |
| longest_run_of_d | the longest repetition of the symbol d |
| longest_run_of_e | the longest repetition of the symbol e |
| longest_run_of_f | the longest repetition of the symbol f |
| longest_run_of_g | the longest repetition of the symbol g |
| longest_run_of_h | the longest repetition of the symbol h |
| longest_run_of_i | the longest repetition of the symbol i |
| longest_run_of_j | the longest repetition of the symbol j |
| longest_run_of_k | the longest repetition of the symbol k |
| fraction_of_a | Fraction of symbol a |
| fraction_of_b | Fraction of symbol b |
| fraction_of_c | Fraction of symbol c |
| fraction_of_d | Fraction of symbol d |
| fraction_of_e | Fraction of symbol e |
| fraction_of f | Fraction of symbol f |
| fraction_of g | Fraction of symbol g |
| fraction_of h | Fraction of symbol h |
| fraction_of i | Fraction of symbol I |
| fraction_of j | Fraction of symbol j |
| fraction_of k | Fraction of symbol k |
| self_transition_probability | The probability of the transition $x \rightarrow x$ |
| lempel_ziv_complexity | Lempel ziv complexity of symbol series |
| recurrence_rate | Recurrence rate of symbol series |
| trapping_time | trapping time of symbol series |
| kolmogorov_complexity | Kolmogorov complexity of symbol series |
| average_symbol_duration | average duration for a symbol to occur in a row |
| symbol_frequency_variability | standard deviation of the symbol frequency distribution |
| symbol_transition_entropy | The entropy of the series of symbol transitions |
| distance_to_healthy | The mean distance of the current 128D point to the healthy centroid |
| angles_to_healthy | The mean current angle ($X_{(t-1)} - X_{(t)}$) to the vector pointing directly to the healthy centroid |

